# Towards a reliable diagnosis of long Covid

**DOI:** 10.1101/2025.09.10.25335169

**Authors:** Alexander D. Franke

## Abstract

**Objectives:** There are multiple definitions of long Covid but none have been externally validated. Reliability analysis can be used as an alternative to identify clinical features that make a reliable diagnosis.

**Methods:** Between October, 2022, and November, 2023, 25 patients were seen consecutively at a general medicine outpatient clinic and diagnosed with long Covid. Demographic details and clinical features were taken from the patients’ notes. The patients’ referring primary care physician was contacted to confirm if the diagnosis of long Covid remained the principal diagnosis or if another diagnosis had replaced it. Unreliability of the diagnosis of long Covid was defined as whether an alternative diagnosis was offered to explain the patient’s symptoms. Bayesian regression analysis was used to estimate hazard ratios for clinical variables and Weibull analysis was used to quantify the “failure” rate and the time needed for 50% of diagnosis to become unreliable.

**Results:** The total follow up time for the cohort was 25.3 patient years. Seventeen of the 25 patients diagnosed with long Covid (72%) were female sex, the median age was 45 years (range, 19 to 78 years), and the median duration of symptoms before the diagnosis of long Covid was 9.9 months (range, 3.4 to 25.4 months). Six patients received an unreliable diagnosis (24%, 95% CrI 11 to 43%). Variables predictive of an unreliable diagnosis were “fatigue”, “headache”; other differential diagnoses plausible after assessment; reduced exercise tolerance; male sex; and age. Variables that were predictive of a reliable diagnosis were "dizziness", palpitations; dyspnoea; myalgia; "memory difficulties" or "memory problems"; "poor concentration" or "brain fog"; migraine; post exercise malaise; patients with more comorbidities; and investigative tests performed after the first assessment. The predicted time needed for 50% of the diagnoses to be unreliable was 892 days.

**Conclusions:** An active diagnostic process, which excluded the existence of alternative diagnoses which could explain a patient’s symptoms, was more likely to produce a reliable diagnosis of long Covid. Reliability analysis can provide clinicians with useful information to improve diagnostic reliability even in circumstances where scientific knowledge of a disease entity is lacking.

**Competing interest statement:** No competing interests

**Funding statement:** No external funding was used

**Author declarations:** I confirm all relevant ethical guidelines have been followed, and any necessary IRB and/or ethics committee approvals have been obtained.

Quality Improvement Medical, General Medicine and Acute Medical Unit Committee of the Fiona Stanley Fremantle Hospital Group gave ethical approval for this work.

## Introduction

Long Covid is a recognized sequala of acute COVID-19 infection, however, there is no universally accepted case definition [1].

Since the publication of the first cohort database by the Patient Led Research Team in 2020 [2], four international health institutions have published four separate definitions of long Covid [3-6] and two organisations have maintained two separate patient cohort databases [7,8]. The NICE [3] definition was followed shortly by the WHO [4], CDC [5], and then the NASEM definitions [6]. The RECOVER consortium have not published a case definition, but have posited a numerical scoring system of symptoms as an alternative way of diagnosing the disease [7]. The REACT group have published longitudinal data on long Covid patients, but have not published a case definition of the disease [8]. None of the existing definitions stipulate qualifying symptoms, but all accept that an infection of COVID-19 precedes the development of long Covid, however, all four differ on the time interval between the acute infection and subsequent development of long Covid. None of these case definitions have been validated in separate cohorts, which leaves the treating clinician unable to confidently say whether any of the promulgated case definitions are suitable. For this reason, it is not currently possible to say if a diagnosis of long Covid is “accurate.” There is also a social function that a given diagnosis performs [9], and in this respect the term “reliable” is more able to convey this social usefulness rather than “accuracy.”

Reliability analysis is concerned with the performance of an object without failure, and the techniques of reliability analysis have been used to quantify medical diagnostic accuracy [10]. Given the lack of a settled definition of long Covid, reliability analysis can be used in place of validation studies to give an empirical measurement of how reliable the diagnosis of long Covid is.

## Methods

Between October, 2022, and November, 2023, 25 patients were seen consecutively at a general medicine outpatient clinic and diagnosed with long Covid. Demographic details and clinical features were taken from the patients’ notes. No patient reported outcome measures were used in the assessment of patients. A symptom qualified as being due to long Covid according to the judgement of the treating physician. The measure of socioeconomic status used was the Index of Relative Socio-economic Disadvantage (IRSD), ranging from 0 to 100, where a lower score indicates greater disadvantage [11]. The Charlson Comorbidity Index (CCI) was calculated using an updated version [12]. Unreliability of the diagnosis of long Covid was defined as whether an alternative diagnosis was offered to explain the patient’s symptoms. A patient’s referring primary care physician was contacted to confirm if the diagnosis of long Covid remained the principal diagnosis or if another diagnosis had replaced it. The Health Department of Western Australia maintains a data linkage system for each patient that has contact with the public health system in the state and this was also checked at follow up. If there was no contact with a health care provider, either the primary care physician or the public health care system, the diagnosis was assumed to be reliable. Quality Improvement Medical, General Medicine and Acute Medical Unit Committee of the Fiona Stanley Fremantle Hospital Group gave ethical approval for this work.

## Statistical plan

Survival analysis for the hazard of an unreliable diagnosis was done using a Cox proportional hazard model which was then used to fit a semiparametric piecewise constant hazard model using a Bayesian regression model. This Bayesian regression model used integrated nested Laplace approximations. Hazard ratios were computed with 95% credible intervals (95% CrI). Single point maximum likelihood estimations (MLEs) were computed with 95% CrI using equal tailed prior probabilities. Point estimates for rate differences were computed with 95% CrI using method of variance estimates recovery. The unpaired Student’s t-test was used for population means. For the Weibull analysis, a two parameter Benard median ranked regression model was used with probability as the dependant variable. These analyses were done using R (version 4.2.1, 2022, https://www.R-project.org/).

## Results

All 25 patients had contact with either their primary health care provider or the public health system in Western Australia at some point during follow up [table 1]. The median time to follow up was 10.6 months (range, 1.6 to 25.5 months) and the total follow up time for the cohort was 25.3 patient years. Seventeen of the 25 patients diagnosed with long Covid (72%) were female sex. The median age was 45 years (range, 19 to 78 years), and the median duration of symptoms before the diagnosis of long Covid was 9.9 months (range, 3.4 to 25.4 months). The median CCI score was zero (range, 0 to 4). Nearly all patients had investigations completed before they were assessed (95%, 95% CrI 83 to 100%). Additional investigations were obtained for 28% (95% CrI 13 to 47%) of the cohort after the first assessment. Long Covid was the only differential for 16 of the patients. For eight patients, long Covid was a differential diagnosis along with one other diagnosis. There were four differential diagnoses entertained for one patient. At the end of the follow up, six patients (24%, 95%CrI 11 to 43%) had an unreliable diagnosis of long Covid [table 2]. Patients with unreliable diagnoses did not differ demographically from those with reliable diagnoses nor were they different in their symptom profiles. Two of the six cases were revised by other physicians, one case of dementia and one case of Parkinsons disease. In three cases, the revised diagnoses were in the original list of differentials after the patient was first assessed. In the remaining instance, the diagnosis of long Covid was no longer preferred by the patient and the alternative diagnosis of small intestinal bacterial overgrowth was substituted.

**Table 1:** Demographic and clinical features of patients diagnosed with long Covid. MLE, maximum likelihood estimation. CrI: credible interval. Quotations indicate patient reported symptoms. †CrI for the difference in rates are calculated using method of variance estimates recovery and there is no statistically significant difference between the two rates if the interval crosses 1; p-values are calculated using the unpaired Student’s t-test, the threshold for statistically significant difference is p<0.05

| Variable | All n=25 | Long Covid reliable n=19 | Long Covid unreliable n=6 | Test for significant difference reliable v. unreliable (95% CrI or p value)† |
| --- | --- | --- | --- | --- |
| Female (n)(MLE%,95%CrI) | 18 (72,53-87) | 14 (73,52-89) | 4 (66,29-92) | -27 - 48 |
| Age (median years)(range) | 45(19-78) | 42(19-74) | 60(40-78) | p=0.2 |
| IRSD (median)(range) | 59(29-98) | 76(29-98) | 52(50-86) | p=0.2 |
| Duration of symptoms (median months)(range) | 9.9(3.4-25.4) | 9.9(3.8-25.4) | 10.5(3.4-15.8) | p=0.5 |
| CCI (median)(range) | 0(0-4) | 0(0-1) | 0(0-4) | p=0.3 |
| Proportion with investigations done prior to the first assessment (MLE%,95%CrI) | 24(95,83-100) | 19(88-100) | 5(81,44-98) | -3 - 55 |
| Proportion with investigations done after the first assessment (MLE%95%,CrI) | 7(28,13-47) | 6(32,14-54) | 1(19,2-56) | -28 - 41 |
| Differential diagnoses per patient at first assessment (median)(range) | 1(1-4) | 1(1-2) | 1(1-4) | p=0.5 |
| Time to follow up/census (median, months)(range) | 10.6(1.6-25.5) | 12.0(2.8-25.5) | 8.4(1.6-18.7) | p=0.4 |
| Symptom or sign (n)(MLE%,95%CrI) | //////// |  |  |  |
| "Fatigue" | 19(76,57-89) | 15(78,57-92) | 4(66,29-92) | -21 - 52 |
| "Poor concentration" or "brain fog" | 12(48,30-67) | 11(58,36-78) | 1(19,2-56) | -4 - 65 |
| Reduced exercise tolerance | 8(32,16-51) | 5(27,11-48) | 3(50,17-83) | -60 - 16 |
| Post exertional malaise | 7(28,13-47) | 6(32,14-54) | 1(19,2-56) | -28 - 41 |
| Dyspnoea | 6(24,11-43) | 5(27,11-48) | 1(19,2-56) | -32 - 36 |
| "Headache" | 6(24,11-43) | 5(27,11-48) | 1(19,2-56) | -32 - 36 |
| Tachycardia | 4(16,6-34) | 3(16,5-36) | 1(19,2-56) | -41 - 24 |
| Myalgia | 4(16,6-34) | 4(22,8-43) | 0(0,0-33) | -15 - 39 |
| Parasthesiae | 3(13,3-29) | 1(6,0.6-22) | 2(34,8-71) | -66 - 3 |
| "Chest pain" | 2(9,2-23) | 2(11,2-30) | 0(0,0-33) | -23 - 26 |
| "Dizziness" | 2(9,2-23) | 2(11,2-30) | 0(0,0-33) | -23 - 26 |
| "Memory difficulties" or "memory" | 2(9,2-23) | 2(11,2-30) | 0(0,0-33) | -23 - 26 |
| problems" |  |  |  |  |
| Migraine | 2(9,2-23) | 2(11,2-30) | 0(0,0-33) | -23 - 26 |
| Palpitations | 2(9,2-23) | 2(11,2-30) | 0(0,0-33) | -23 - 26 |
| Postural hypotension | 2(9,2-23) | 1(6,0.6-22) | 1(19,2-56) | -50 - 11 |
| Agitation | 1(5,0.4-17) | 0(0,0-12) | 1(19,2-56) | -55 - 3 |
| "Anxiety" | 1(5,0.4-17) | 1(6,0.6-22) | 0(0,0-33) | -27 - 19 |
| Arthralgia | 1(5,0.4-17) | 1(6,0.6-22) | 0(0,0-33) | -27 - 19 |
| "Back pain" | 1(5,0.4-17) | 0(0,0-12) | 1(19,2-56) | -55 - 3 |
| "Bodily weakness" | 1(5,0.4-17) | 1(6,0.6-22) | 0(0,0-33) | -27 - 19 |
| "Body pain" | 1(5,0.4-17) | 1(6,0.6-22) | 0(0,0-33) | -27 - 19 |
| Cough | 1(5,0.4-17) | 1(6,0.6-22) | 0(0,0-33) | -27 - 19 |
| "Insomnia" | 1(5,0.4-17) | 1(6,0.6-22) | 0(0,0-33) | -27 - 19 |
| "Internal tremor" | 1(5,0.4-17) | 1(6,0.6-22) | 0(0,0-33) | -27 - 19 |
| "Internal vibration" | 1(5,0.4-17) | 1(6,0.6-22) | 0(0,0-33) | -27 - 19 |
| "Muscle weakness" | 1(5,0.4-17) | 1(6,0.6-22) | 0(0,0-33) | -27 - 19 |
| "Night sweats" | 1(5,0.4-17) | 1(6,0.6-22) | 0(0,0-33) | -27 - 19 |
| "Pain" | 1(5,0.4-17) | 1(6,0.6-22) | 0(0,0-33) | -27 - 19 |
| Panic attacks | 1(5,0.4-17) | 0(0,0-12) | 1(19,2-56) | -55 - 3 |
| Pre-syncope | 1(5,0.4-17) | 1(6,0.6-22) | 0(0,0-33) | -27 - 19 |
| "Shakiness" | 1(5,0.4-17) | 1(6,0.6-22) | 0(0,0-33) | -27 - 19 |
| Soft neurological signs | 1(5,0.4-17) | 1(6,0.6-22) | 0(0,0-33) | -27 - 19 |
| Throat infections | 1(5,0.4-17) | 1(6,0.6-22) | 0(0,0-33) | -27 - 19 |
| Tinnitus | 1(5,0.4-17) | 1(6,0.6-22) | 0(0,0-33) | -27 - 19 |
| Unilateral neurological symptoms | 1(5,0.4-17) | 0(0,0-12) | 1(19,2-56) | -55 - 3 |
| Vertigo | 1(5,0.4-17) | 0(0,0-12) | 1(19,2-56) | -55 - 3 |
| Weight loss | 1(5,0.4-17) | 0(0,0-12) | 1(19,2-56) | -55 - 3 |
| "Wheeze" | 1(5,0.4-17) | 0(0,0-12) | 1(19,2-56) | -55 - 3 |

**Table 2:** Clinical features of the cases where the diagnosis of long Covid was unreliable.

| Unreliable diagnosis | Sex | Age range (years) | C CI | IRS D | Duration of symptoms (months) | Investigative tests before first assessment | Investigative tests after first assessment | Follow up time (months) | Symptoms and signs | Differential diagnoses at the time of first assessment | Diagnosis at census |
| --- | --- | --- | --- | --- | --- | --- | --- | --- | --- | --- | --- |
| 1 | male | 76-80 | 4 | 52 | 8.7 | yes | no | 4.2 | Fatigue<br>Post exertional malaise<br>Parasthesiae<br>Reduced exercise tolerance | none | Dementia |
| 2 | female | 66-70 | 0 | 57 | 9.0 | yes | no | 1.6 | Fatigue<br>Unilateral | none | Parkinsons |
|  |  |  |  |  |  |  |  |  | al<br>neurolo<br>gical<br>sympto<br>ms<br>Agitatio<br>n<br>Soft<br>neurolo<br>gical<br>signs |  | disease |
| 3 | fem<br>ale | 76-80 | 1 | 50 | 3.4 | yes | no | 6.2 | Parasthe<br>siae<br>Postural<br>hypoten<br>sion | 1)Postura<br>l<br>hypotens<br>ion | Postural<br>hypoten<br>sion |
| 4 | fem<br>ale | 31-35 | 0 | 52 | 13.3 | yes | yes | 18.7 | Fatigue<br>Dyspno<br>ea<br>Tachyca<br>rdia<br>Reduced<br>exercise<br>toleranc<br>e | 1)3<sup>rd</sup><br>trimester<br>2)Pulmo<br>nary<br>hyperten<br>sion<br>3)Obesit<br>y | Obesity |
| 5 | fem<br>ale | 51-55 | 0 | 52 | 12.0 | no | no | 18.0 | Reduced<br>exercise<br>toleranc<br>e<br>Panic<br>attacks<br>Wheeze | 1)Menop<br>ause | Menopa<br>use |
| 6 | mal<br>e | 36-40 | 0 | 86 | 15.8 | yes | no | 10.6 | Fatigue<br>Brain<br>fog<br>Headach<br>e<br>Back<br>pain<br>Vertigo<br>Weight<br>loss | none | Small<br>intestina<br>l<br>bacterial<br>overgro<br>wth |

Patient reported outcome measures were not used. For instance, though there are quantitative outcome measures for symptoms like “memory difficulties” or “fatigue” only patients declaring them as troublesome was required to qualify as a symptom of long Covid.

Patients were asked to clarify their symptoms as related to exercise. Post exertional malaise was present if a worsening of symptoms developed hours or days after an activity, was disproportionate to the activity, and had a prolonged recovery phase. Reduced exercise tolerance occurred when a patient was unable to sustain their normal effort while completing an activity. Tachycardia and weight loss were either documented on the referral to the clinic or found on physical examination. Dyspnoea, myalgia, parasthesiae, migraine, palpitations, postural hypotension, agitation, arthralgia, cough, panic attacks, pre-syncope, tinnitus, and vertigo were present if they fit the received definitions. The one case of recurrent throat infections was documented in the referral. One patient had unilateral neurological symptoms of stiffness and tremor and had soft neurological signs on physical examination. Of the 38 symptoms and signs, 15 were possessed by two or more patients [table 1]. The most common symptoms were fatigue (76%, 95% CrI 57 to 89), “poor concentration” or “brain fog” (48%, 95% CrI 30 to 67%); reduced exercise tolerance (32%, 95% CrI 16 to 51%); post exertional malaise (28%, 95% CrI 13 to 47%); dyspnoea (24%, 95% CrI 11 to 43%); “headache” (24%, 95% CrI 11 to 43%); tachycardia (16%, 95% CrI 6 to 34%); myalgia (16%, 95% CrI 6 to 34%); and parasthesiae (13%, 95% CrI 3 to 29%). Each of the symptoms and signs of “chest pain”, “dizziness”, “memory difficulties” or “memory problems”, migraine, palpitations, and postural hypotension occurred in 9% (95% CrI 2 to 23%) of patients, a piece.

Symptoms and clinical signs were included as variables in the hazard model if at least two patients possessed them [figure 1]. The variables for which the HR 95% CrI included one, indicating they were not predictive of either an unreliable or reliable diagnosis of long Covid, were less socioeconomic disadvantage, longer duration of symptoms; investigative tests performed before the first assessment; tachycardia; parasthesiae; “chest pain”; and postural hypotension. The variables for which the HR 95% CrI remained below one, indicating that the variable was predictive of a reliable diagnosis of long Covid, were higher CCI, investigative tests performed after the first assessment; “poor concentration” or “brain fog”; post exercise malaise; dyspnoea; myalgia; “memory difficulties” or “memory problems”; palpitations; “dizziness”; and migraine. The variables for which the HR 95% CrI remained above one, indicating that the variable was predictive of an unreliable diagnosis of long Covid, were male sex, higher age; other differential diagnoses considered plausible; “fatigue”; reduced exercise tolerance; and “headache.”

**Figure 1.**
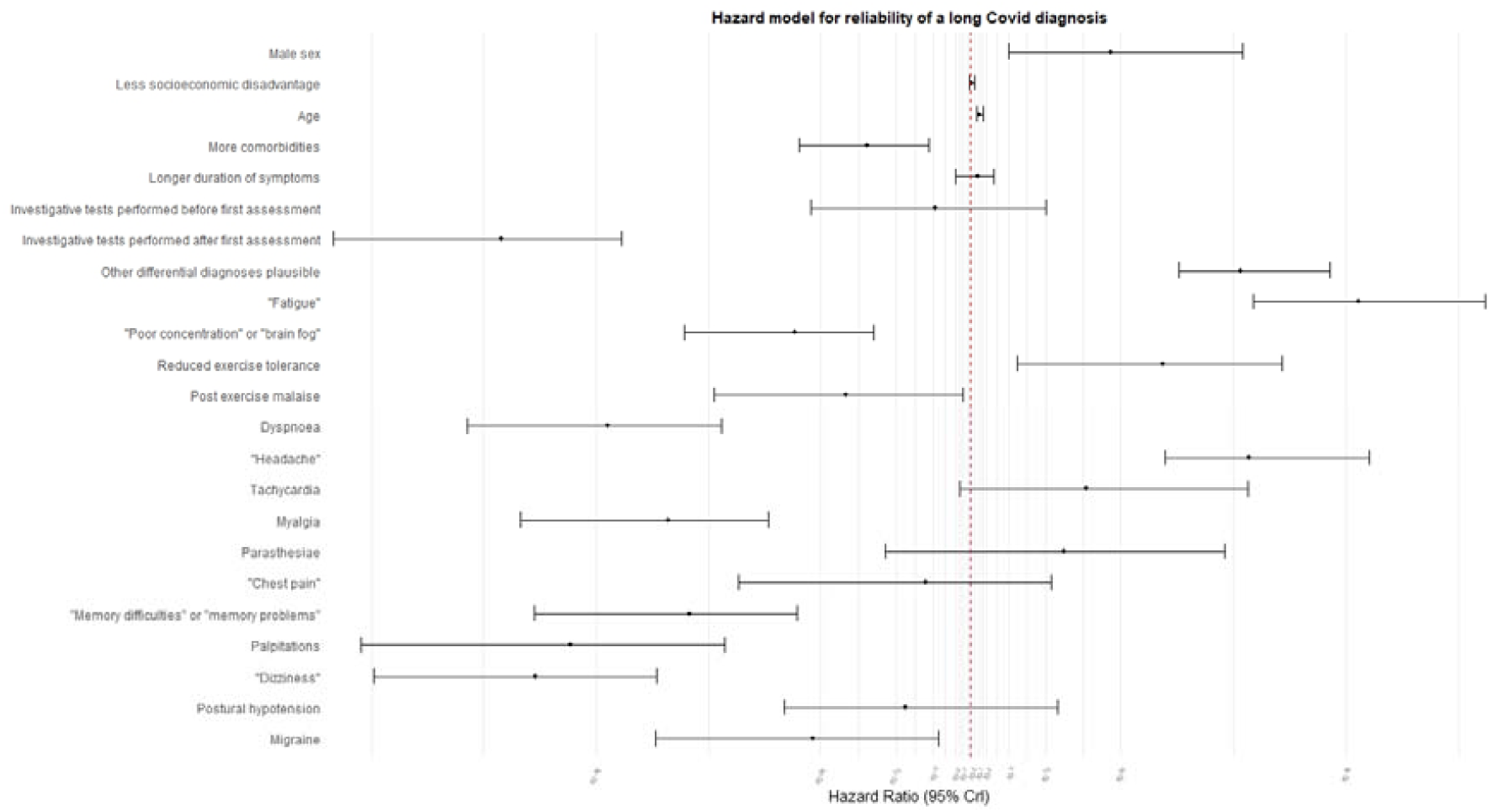
Forest plot for clinical variables predictive of a reliable diagnosis of long Covid.

Figure 2 shows the Weibull log-log plot of the number of unreliable long Covid diagnoses over time and figure 3 shows the cumulative probability for an unreliable diagnosis. The “failure rate”, or in the present case the rate of unreliable diagnoses, is calculated by estimating the slope of the line of best fit for a log-log plot. This is also known as the shape parameter (β). When β is less than 1, the “failure rate” declines over time; if β is equal to 1, the rate is described as “random failure”; and if β is more than 1, the “failure rate” increases with time. A high Pearson correlation coefficient indicates a strong relationship between the observed “failure rate” and the line of best fit.

**Figure 2.**
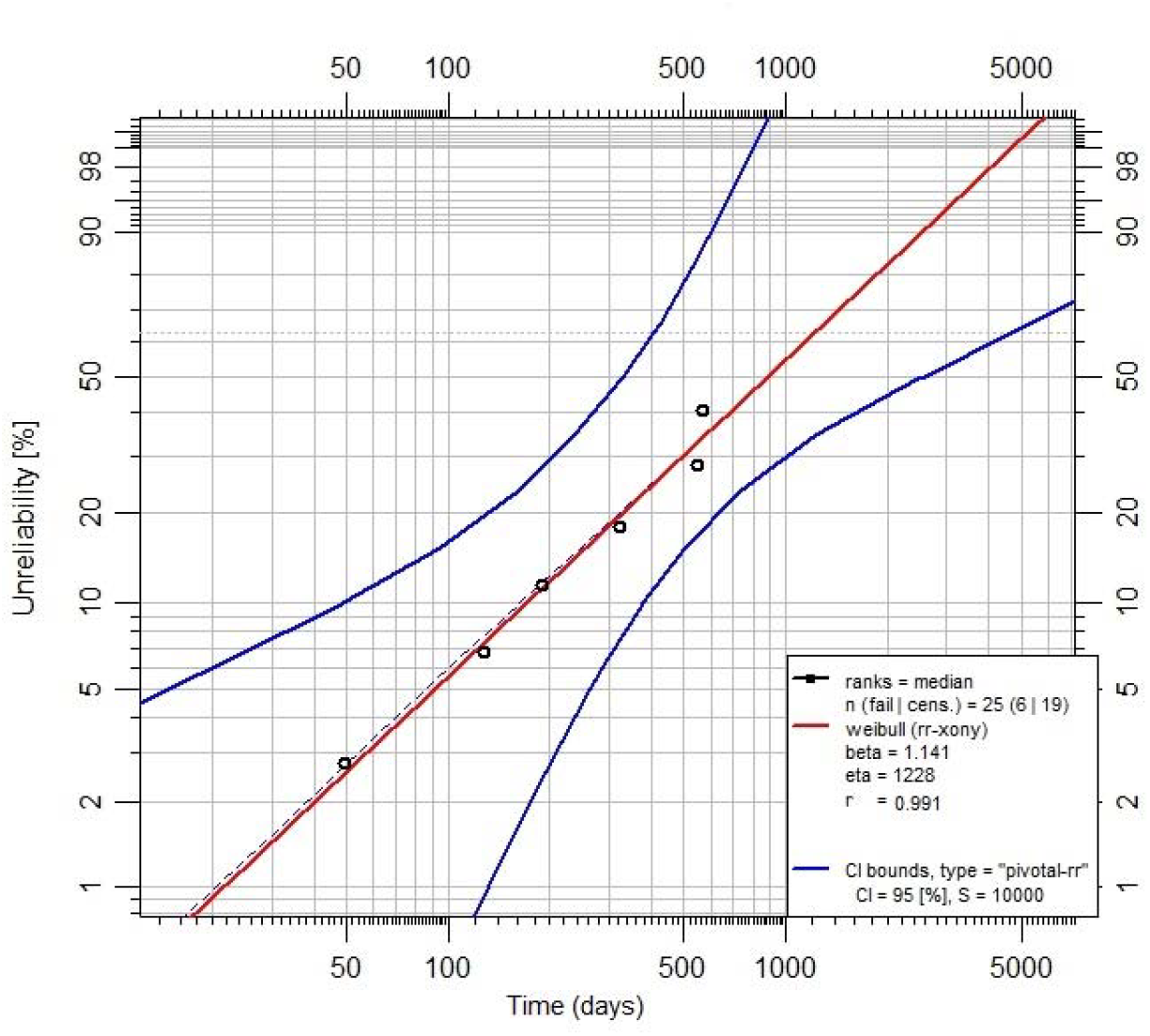
Weibull log-log plot of the number of unreliable long Covid diagnoses

**Figure 3.**
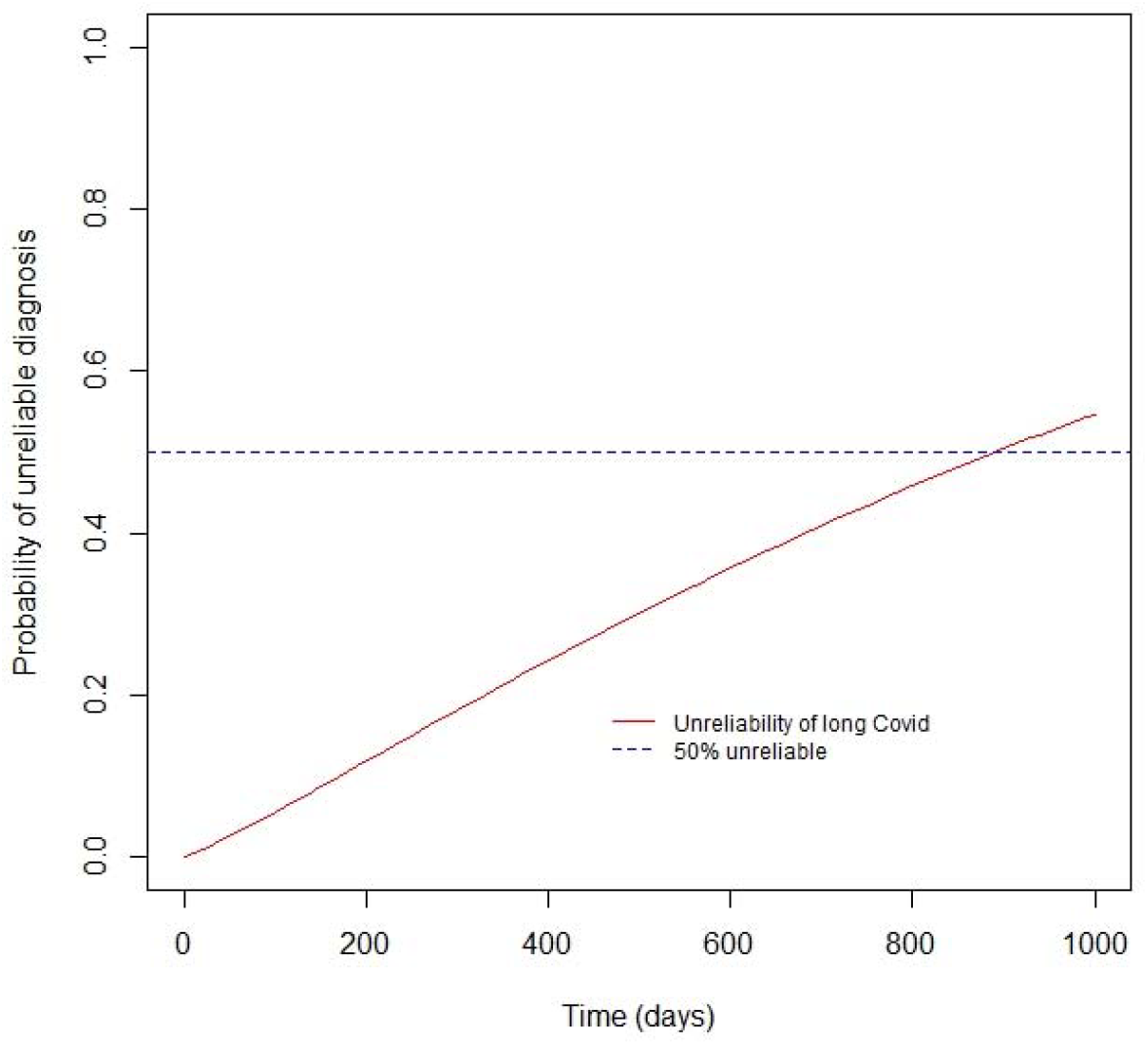
Cumulative probability of an unreliable diagnosis of long Covid.

The “failure rate” for a diagnosis of long Covid was 1.14, which meant the rate of unreliable diagnoses of long Covid increased over time. The predicted time for unreliable diagnoses to reach 50% was 892 days and the correlation coefficient was 0.99.

## Discussion

Each of the 25 patients in the present cohort would have qualified under all current definitions of long Covid. All had a history where the initial Covid infection was either probable or confirmed by testing. All had symptoms for at least three months, and all experienced a functional deficit from their symptoms. After 25.3 patient-years of follow up, six patients were found to have an unreliable diagnosis of long Covid. There were no between group differences in demographic variables, nor was there a preponderance of a given symptom in one group compared to the other. The variables that predicted a reliable diagnosis of long Covid were higher CCI, investigative tests performed after the first assessment; “poor concentration” or “brain fog”; post exercise malaise; dyspnoea; myalgia; “memory difficulties” or “memory problems”; palpitations; “dizziness”; and migraine. The variables that predicted an unreliable diagnosis were male sex, higher age; other differential diagnoses considered plausible; “fatigue”; reduced exercise tolerance; and “headache.” According to the reliability analysis, unreliability increased with time, and the time taken for 50% of diagnoses to be unreliable was predicted to be approximately 890 days. Given that a reliable diagnosis was more likely if investigative tests were performed after the first assessment and an unreliable diagnosis was more likely if there were other differential diagnoses entertained at the first assessment, the increasing unreliability with time reflects that long Covid was a diagnosis made by exclusion.

Traditionally, organizations, usually professional bodies, devise diagnostic criteria for a particular disease, and some of these criteria subsequently have formal validation studies to quantify their sensitivity and specificity for the disease. Applying reliability analysis to medical diagnostic decision making adds the unique ability to quantify “failure rate” of a diagnosis as well as give a predicted time for a given percentage of diagnoses to “fail”. Using these two parameters it is possible to compare diagnostic techniques and is therefore an alternative to the traditional method of quantifying sensitivity and specificity for a diagnostic instrument.

The WHO and NICE diagnostic criteria both state that symptoms of long Covid must not be explained by “alternative diagnoses” [3,4]. The NASEM definition takes the opposite approach by listing multiple separate diagnoses that can be considered as synonymous with long Covid [6]. The possibility of alternative diagnoses when the patient was first assessed indicated an unreliable diagnosis, and diagnostic tests that were obtained after the patient’s first assessment, not before it, was indicative of a reliable diagnosis. These could be considered as proxies for an active diagnostic process applied to the presentation of patients complaining of symptoms due to long Covid, rather than an approach that equates long Covid with a myriad of other disease entities.

This report highlights the difficulties in identifying the lead symptoms for the diagnosis of long Covid. “Poor concentration or brain fog” and post exercise malaise were both indicators of a reliable diagnosis in the present cohort. The RECOVER and WHO cohorts reported “brain fog” in 64% and 74% of respondents, respectively, while the Patient Led Research Team reported a prevalence of 24%; it was absent in the REACT cohort. Of the international cohorts, only RECOVER reported post exercise malaise. Symptoms related to memory, indicative of a reliable diagnosis in this cohort, were only reported in the WHO cohort. For the other symptoms that were indicative of a reliable diagnosis, the prevalence ranged from 9% [8] to 78% [4] for dyspnoea, 6% [8] to 64% [4] for myalgia; and 3% [8] to 47% [4] for dizziness. Palpitations were reported in the WHO (60%) and RECOVER (57%) cohorts. No other cohort reported migraine as a prevailing symptom. In the present cohort, “fatigue” was found to indicate an unreliable diagnosis of long Covid. The first report of long Covid, by the Patient-Led Research Team, cited a prevalence of “mild or moderate fatigue” of 65%, eight weeks after Covid infection, while “tiredness” was present for 15% of respondents in the REACT-2 cohort. “Headache”, too, was an indicator of an unreliable diagnosis in the present cohort, and similarly the prevalence varied widely in the other cohorts, ranging from 5% to 56% in the REACT and WHO cohorts, respectively. The lack of a universal definition for “fatigue” and “headache” applied to all cohorts may account for this discordancy. None of the other cohorts reported on reduced exercise tolerance, which was found to indicate an unreliable diagnosis in the present cohort.

The external validity of this report is hampered by the small cohort size. Also, as is the case for all long Covid cohorts internationally, the lack of conforming definitions for each symptom or formal patient reported outcome measures makes the comparisons with external cohorts challenging.

In conclusion, reliability analysis was able to identifying demographic features, symptoms, and clinical signs that predicted a reliable diagnosis of long Covid. An active diagnostic process, which excluded the existence of alternative diagnoses which could explain a patient’s symptoms, was more likely to produce a reliable diagnosis of long Covid. Reliability analysis can provide clinicians with useful information to improve diagnostic reliability even in circumstances when scientific knowledge of a disease entity is lacking.

## Data Availability

All data produced in the present study are available upon reasonable request to the authors

